# An updated PREDICT breast cancer prognostic model including the benefits and harms of radiotherapy

**DOI:** 10.1101/2023.07.18.23292777

**Authors:** Isabelle Grootes, Gordon C. Wishart, Paul David Peter Pharoah

## Abstract

**Background:** Predict Breast (www.breast.predict.nhs.uk) is an online prognostication and treatment benefit tool for early invasive breast cancer. However, the most recent version of PREDICT Breast (v2.2) was based on data for breast cancer cases diagnosed from 1999 to 2003 and did not incorporate the benefits of radiotherapy or the harms associated with theray. Since then, there has been a substantial improvement in the outcomes for breast cancer cases. The aim of this study was to update PREDICT Breast to ensure that the underlying model is appropriate for contemporary patients.

**Methods:** Data from 4,644 ER-negative and 30,830 ER-positive breast cancer cases diagnosed from 2000 to 2017 in the region served by the Eastern Cancer Registry were used for model development. Multivariable fractional polynomials in a Cox proportional hazards framework were used to estimate the prognostic effects of year of diagnosis, age at diagnosis, tumour size, tumour grade and number of positive nodes and to compute the baseline hazard functions. Separate models were developed for ER-positive and ER-negative disease. Data on 32,408 breast cancer patients from the West Midlands Cancer Registry and from 100,551 breast cancer cases from the other English Cancer Registries combined were used to determine the discriminative power, calibration, and reclassification of the new version of PREDICT Breast (v3.0).

**Results:** The new model (v3.0) was well-calibrated; predicted numbers of 5-, 10- and 15-year breast cancer deaths were within 10 per cent of the observed number in both model development and model validation data sets. In contrast, PREDICT Breast v2.2 was found to substantially over-predict the number of deaths. Discrimination was also good: The AUC for 15-year breast cancer survival was 0. 824 in the model development data, 0.809 in the West Midlands data set and 0.846 in the data set for the other registries. There figures were slightly better than those for PREDICT Breast v2.2

**Conclusion:** Incorporating the prognostic effect of year of diagnosis, updating the prognostic effects of all risk factors and amending the baseline hazard functions have led to an improvement of model performance of PREDICT Breast. The new model will be implemented in the online tool which should lead to more accurate absolute treatment benefit predictions for individual patients.

## INTRODUCTION

The PREDICT breast cancer prognostication and treatment benefit prediction model (v1) was developed in 2010 using data from the UK East Anglia Cancer Registration and Information Centre (ECRIC) for model fitting and data from the West Midlands Cancer Intelligence Unit for model validation ^1-3^. The model fitting data set comprised data on 5,232 cases diagnosed from 1999 to 2003. PREDICT v1 was implemented as a web-based tool for clinicians in January 2011 (www.breast.predict.nhs.uk), and since then the use of the tool has increased steadily around the world. The model was refitted in 2017 using the original cohort of cases from East Anglia with updated survival time in order to take into account age at diagnosis and to smooth out the hazard ratio functions for tumour size and node status (v2) ^4^. PREDICT has been independently validated in cohorts from Canada ^5^, Malaysia ^6^, the Netherlands ^7-9^, and the UK ^10 11^ and has generally been shown to have good discrimination and calibration.

The data on which PREDICT breast v1 and v2 was based were breast cancer cases diagnosed in the Eastern Region of England over 20 years ago. Since then, the prognosis of early breast cancer has improved substantially ^12^ and it is likely that the current model is not well calibrated for contemporary patients ^13^. Moreover, the number of cases with ER negative disease in the cohort was comparatively small (<1,000) and it is possible that the estimates of the prognostic effects of the variables in the ER negative disease model were sub-optimal. Furthermore, radiotherapy and chemotherapy have been shown to associated with an increase in mortality from causes other than breast cancer ^14 15^ and this was not taken into account in previous versions of PREDICT Breast

We have therefore refitted the PREDICT breast model using a national data set of patients diagnosed from 2000 to 2017 with the aim of refining the hazard ratio estimates for the variables in the current model and to estimate the effect of year of diagnosis on prognosis in order to be able to recalibrate the model for contemporary patients. In addition, we included the beneficial effect of radiotherapy on breast cancer mortality and the harmful effect of both chemotherapy and radiotherapy of other causes of mortality. Model development, validation and reporting were carried out according to the TRIPOD (Transparent Reporting of a multivariable prediction model for Individual Prognosis Or Diagnosis) criteria ^16^.

## METHODS

### Patient data

Public Health England provided data from the National Cancer Registration and Analysis Service (PHE NCRAS) for all women diagnosed with non-metastatic breast cancer from 2000 to 2017 inclusive. Information obtained from PHE NCRAS included age at diagnosis, year of diagnosis, tumour size, histological grade, tumour stage at diagnosis, number of lymph nodes sampled, number of lymph nodes positive, ER status, HER2 status, mode of detection (clinically detected vs. screen detected), and whether the patient had undergone chemotherapy, hormone therapy and/or radiotherapy for two time periods, the first being within 6 months following their diagnosis and the second being treatments received throughout their entire follow-up time. Patients younger than 25 or older than 85 at diagnosis, patients with a tumour larger than 20 centimetres, or with more than 20 positive lymph nodes were excluded from the analysis. Of 372,110 cases, complete data were available for 163,224 (44%). Initial analyses showed that the Eastern Cancer Registry and the West Midlands Cancer Registry had fewer missing data (62% and 71% complete cases) compared to the other registries (35% complete cases) particularly in years 2000 to 2009 (Supplementary Table 1). The variable with the most missing data was ER status (42% missing), 31% were missing number of positive nodes, 16% were missing tumour size, 3% were missing tumour grade and 6% were missing mode of detection. The complete case data set for the Eastern Cancer Registry (n = 35,474; 4,644 ER-negative and 30,830 ER-positive) was used for the development of the new version of PREDICT Breast and the West Midlands Cancer Registry data set (n = 31,801; 4,668 ER-negative; 27,133 ER-positive) was used as the primary validation data and the data set for the other cancer registries (n = 95,949; 12,814 ER-negative; 83,135 ER-positive) used as an additional validation data set.

Details of the specific regimen used for chemotherapy were not available and we assumed that all patients that underwent chemotherapy were treated with an anthracycline-based regimen. Nor was information on trastuzumab and bisphosphonate therapy available. The benefits of radiotherapy were applied to all patients who received including those who had lumpectomy and those who had mastectomy as the primary surgical treatment. Death certificate flagging through the Office for National Statistics provides the registries with notification of deaths. The lag times for these are a few weeks for cancer deaths and 2 months to 1 year for non-cancer deaths. Vital status was ascertained at the end of December 2019, and so all analyses were censored on 31 December 2018 to allow for delay in reporting of vital status. Breast cancer-specific mortality was defined as deaths where breast cancer was listed as the cause of death on part 1a, 1b or 1c of the death certificate.

### Statistical methods

Multivariable Cox proportional hazards models were used to estimate the prognostic effect of each variable. In all models follow up time was defined as the time from breast cancer diagnosis to last follow up, death or 15 years after diagnosis, whichever came first. The outcome of interest was either breast cancer-specific mortality or mortality from other causes.

Separate models were derived for breast cancer-specific mortality in ER-negative and ER-positive cases. Multiple fractional polynomials were used to model non-linear effects between the continuous risk factors (age at diagnosis, tumour size and number of positive nodes) and breast cancer-specific mortality as adding higher order polynomials to the model will improve the fit to the data in the presence of non-linearity. Sequential backward elimination with a maximum of 4 degrees of freedom for a single continuous predictor was used to estimate the continuous variable transformations. In addition to the variables already present in the current version of PREDICT, the year of breast cancer diagnosis and the effect of radiotherapy were also incorporated into the analyses. Age at diagnosis was transformed to age at diagnosis minus 24 and year of diagnosis was transformed to year minus 2000 in order that the baseline hazard would be more realistic. The baseline hazard is the hazard that corresponds to a hypothetical individual with all variables taking a value of zero. Transforming age at diagnosis and year at diagnosis in this way means that the baseline hazard corresponds to a woman diagnosed at age 24 in the year 2000 rather than a woman diagnosed at age 0 in the year 0. The relative treatment benefits for chemotherapy, hormone therapy and radiotherapy were constrained to the estimates of benefit randomised controlled trial meta-analyses of the Early Breast Cancer Trialists Collaborative Group (adjuvant hormone therapy log hazard ratio -0.386 ^17^, adjuvant chemotherapy log hazard ratio -0.248 ^18^, radiotherapy log hazard ratio -0.180 ^19^) by adding them as an offset in the analyses. After fitting the Cox proportional hazards models to ER-negative and ER-positive cases, a multiple fractional polynomial model with a Gaussian distribution was fit to the baseline hazards according to the method of Sauberei and colleagues ^20^ to derive a smoothed baseline hazard functions for breast cancer-specific mortality.

A single multivariate Cox regression model for mortality from other causes (non-breast cancer-specific) was built for ER-negative and ER-positive cases combined with year of diagnosis and age at diagnosis modelled using multivariable fractional polynomials. The relative harms of chemotherapy and radiotherapy were constrained to the estimates of benefit reported by Kerr and colleagues (adjuvant chemotherapy log hazard ratio 0.183) ^14^ and Taylor and colleagues (radiotherapy log hazard ratio 0.078 per Gray whole-heart dose) ^15^ by adding them as an offset in the analyses. We assumed all patients receiving radiotherapy receive a whole heart dose of 2 Gy, as radiotherapy dose was not available in our data. The smoothed baseline hazard function for non-breast cancer specific mortality was also computed using a multivariable fractional polynomial model.

### Model validation

The models derived from the Eastern Cancer Registry were used to predict the probabilities of death from breast cancer or death from other causes in the cases in both validation data sets. Because the web version of PREDICT Breast v2.2 allows for missing data on mode of detection we also included 9,848 cases for whom only modes of detection was missing. Model calibration was performed by comparing the observed number of deaths with those predicted by v3.0 and v2.2 up to 5 years, 10 years and 15 years after diagnosis. Calibration plots were used to visualise calibration at different levels of risk. Model discrimination was evaluated by calculating the area under the receiver operator-characteristic curve (AUC) for up to 5-year, 10-year and 15-year breast cancer mortality. The AUC is the probability that the predicted mortality from a randomly selected patient who died will be higher than the predicted mortality from a randomly selected survivor.

All analyses were carried out using the *mfp* ^21^, *patchwork* ^22^, *pROC*, ^23^ *survival* ^24^, *tableone* ^25^ and *tidyverse* ^26^ packages for the R software ^27^ implemented in R Studio ^28^.

## RESULTS

Table 1 shows the patient characteristics by cancer registry. The model fitting was carried out using Eastern Cancer Registry data for 4,644 women with an ER-negative tumour and 34,265 women with an ER-positive tumour.

**Table 1:**
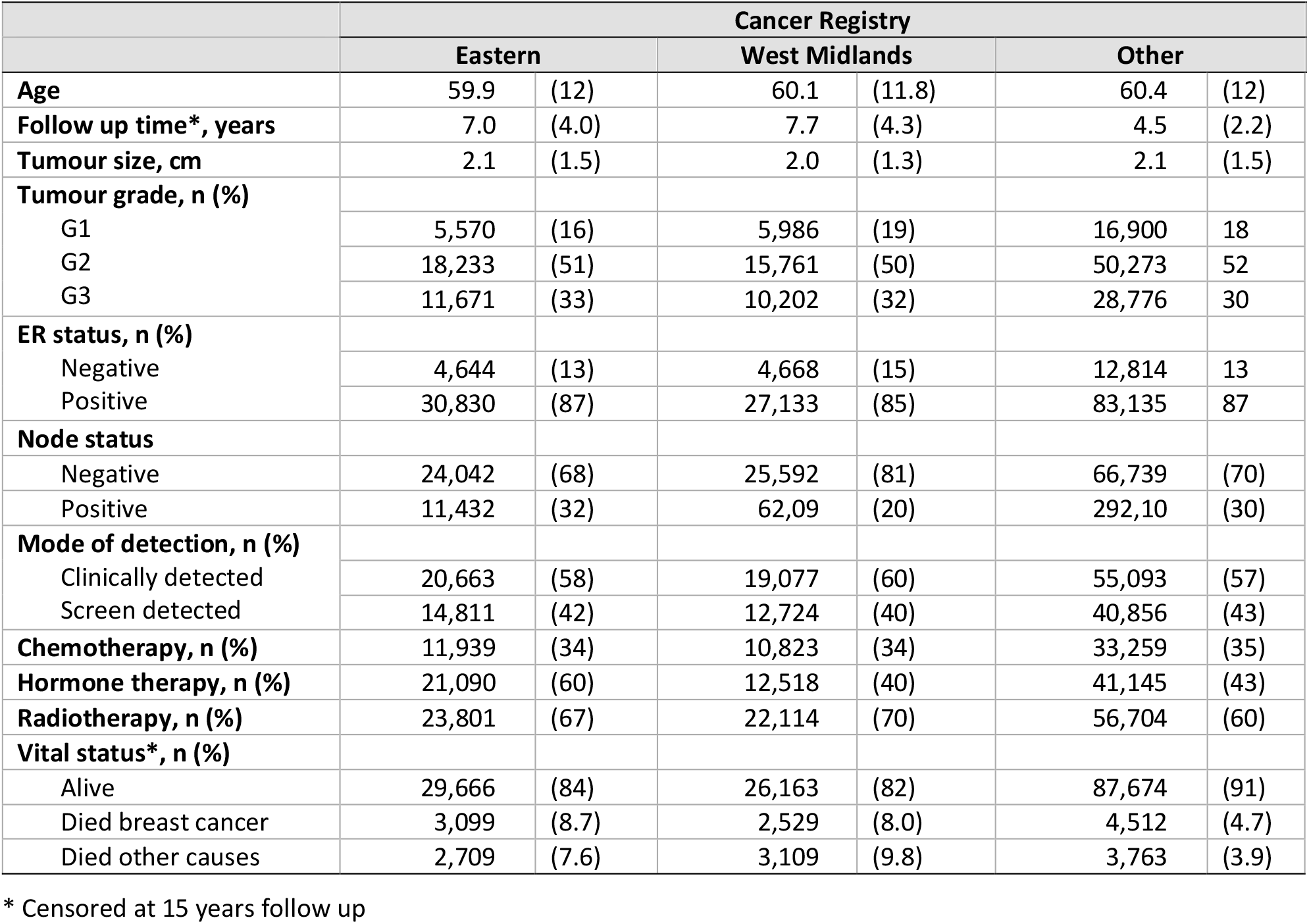
Patient characteristics for the Eastern Cancer Registry, the West Midlands cancer registry and the other cancer registries. Mean (sd), unless stated otherwise.

|  | Cancer Registry |  |  |  |  |  |
| --- | --- | --- | --- | --- | --- | --- |
|  | Eastern |  | West Midlands |  | Other |  |
| **Age** | 59.9 | (12) | 60.1 | (11.8) | 60.4 | (12) |
| **Follow up time\*, years** | 7.0 | (4.0) | 7.7 | (4.3) | 4.5 | (2.2) |
| **Tumour size, cm** | 2.1 | (1.5) | 2.0 | (1.3) | 2.1 | (1.5) |
| **Tumour grade, n (%)** |  |  |  |  |  |  |
| G1 | 5,570 | (16) | 5,986 | (19) | 16,900 | 18 |
| G2 | 18,233 | (51) | 15,761 | (50) | 50,273 | 52 |
| G3 | 11,671 | (33) | 10,202 | (32) | 28,776 | 30 |
| **ER status, n (%)** |  |  |  |  |  |  |
| Negative | 4,644 | (13) | 4,668 | (15) | 12,814 | 13 |
| Positive | 30,830 | (87) | 27,133 | (85) | 83,135 | 87 |
| **Node status** |  |  |  |  |  |  |
| Negative | 24,042 | (68) | 25,592 | (81) | 66,739 | (70) |
| Positive | 11,432 | (32) | 62,09 | (20) | 292,10 | (30) |
| **Mode of detection, n (%)** |  |  |  |  |  |  |
| Clinically detected | 20,663 | (58) | 19,077 | (60) | 55,093 | (57) |
| Screen detected | 14,811 | (42) | 12,724 | (40) | 40,856 | (43) |
| **Chemotherapy, n (%)** | 11,939 | (34) | 10,823 | (34) | 33,259 | (35) |
| **Hormone therapy, n (%)** | 21,090 | (60) | 12,518 | (40) | 41,145 | (43) |
| **Radiotherapy, n (%)** | 23,801 | (67) | 22,114 | (70) | 56,704 | (60) |
| **Vital status\*, n (%)** |  |  |  |  |  |  |
| Alive | 29,666 | (84) | 26,163 | (82) | 87,674 | (91) |
| Died breast cancer | 3,099 | (8.7) | 2,529 | (8.0) | 4,512 | (4.7) |
| Died other causes | 2,709 | (7.6) | 3,109 | (9.8) | 3,763 | (3.9) |

\* Censored at 15 years follow up

On fitting the multivariable fractional polynomial model to the ER-positive cases the hazard ratio function for tumour size was found to be *2*.*39*(size)*^*0*.*5*^ *– 0*.*439*size*. Under this function the hazard ratio would increase to a maximum for a tumour of 7.4 cm and then decrease for larger tumours (Figure 1 dashed line). It seems unlikely that the true effect size would get smaller with increasing tumour size and so we refitted the model using *1 – exp(-size/2)* so that the hazard ratio increases up to 7.5 cm and then flattens off (Figure 1 solid line). The breast cancer-specific mortality hazard ratio (HR) functions for age at diagnosis, tumour size and number of positive nodes for the ER-negative and ER-positive cases are shown in Figure 2 and the associated logarithmic hazard ratios in Table 2.

**Table 2:** Fractional polynomial functions and associated logarithmic hazard ratios for age at diagnosis, tumour size, number of positive nodes, tumour grade and mode of detection by oestrogen receptor (ER) status.

| Prognostic factor | Function | Log HR | p-value |
| --- | --- | --- | --- |
| *ER-negative breast cancer specific mortality* |  |  |  |
| Age at diagnosis 1 | ((age-24)/100) | 1.756 | <0.0001 |
| Age at diagnosis 2 | ((age-24)/100)*log((age-24)/100)) | 4.555 | <0.0001 |
| Tumour size, cm | log(size) | 0.744 | <0.0001 |
| No. positive lymph nodes | log(nodes+1) | 0.631 | <0.0001 |
| Tumour grade | grade - 1 | 0.346 | <0.0001 |
| Mode of detection | screen detected | -0.211 | 0.037 |
| Year of diagnosis | year - 2000 | -0.046 | <0.0001 |
| *ER-positive breast cancer specific mortality* |  |  |  |
| Age at diagnosis 1 | ((age-24)/100)<sup>-0.5</sup> | 0.196 | 0.0004 |
| Age at diagnosis 2 | ((age-24)/100)<sup>2</sup> | 2.929 | <0.0001 |
| Tumour size 1, cm | 1 – exp(-size/20) | 2.274 | <0.0001 |
| No. positive lymph nodes | log(nodes + 1) | 0.672 | <0.0001 |
| Tumour grade | grade - 1 | 0.705 | <0.0001 |
| Mode of detection | screen detected | -0.320 | <0.0001 |
| Year of diagnosis | year | -0.048 | <0.0001 |
| *All cases non breast cancer mortality* |  |  |  |
| Age at diagnosis 1 | ((age-24)/100)<sup>3</sup> | 4.21 | 0.0007 |
| Age at diagnosis 2 | ((age-24)/100)<sup>3</sup>*log((age-24)/100)) | -31.4 | <0.0001 |
| Year of diagnosis | year | -0.021 | 0.0001 |

**Figure 1:**
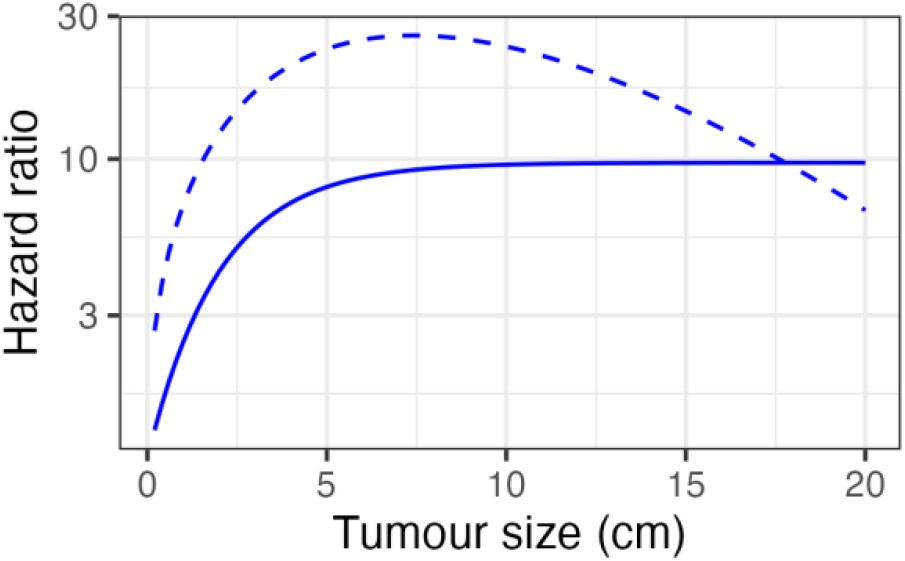
Polynomial hazard ratio functions for tumour size. Dashed line - best fit from multivariable fractional polynomial model. Solid line - monotonic function selected for inclusion in the final model

**Figure 2:**
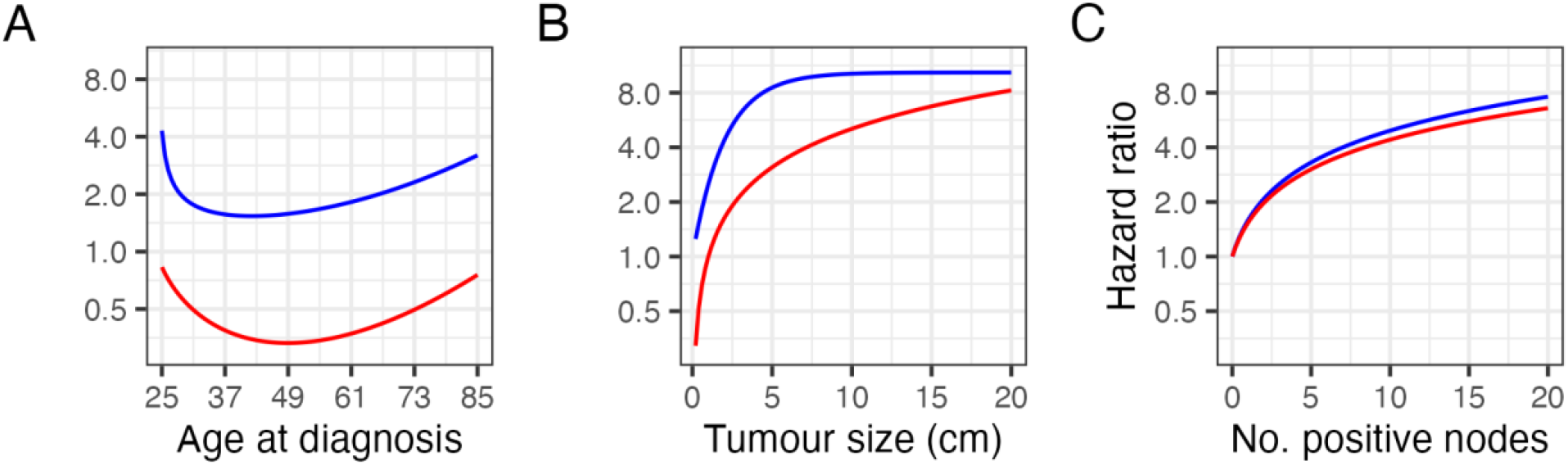
Breast cancer-specific mortality hazard ratio functions. A age, B tumour size and C the number of positive nodes. ER-negative is indicated by red lines and ER-positive is indicated by blue lines.

The derived polynomial baseline hazard functions for breast cancer specific mortality in the ER-negative cases and ER-positive cases and non-breast cancer mortality are given by the following equations:

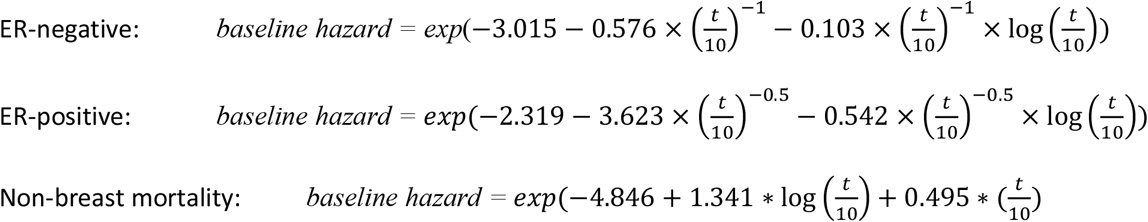

These functions provided a very good fit to the observed baseline hazard (Supplementary Figure 1).

### Model calibration

Table 3 shows the cumulative number of breast cancer deaths predicted at five, ten and 15 years by the new version of the model (v3.0) and the current version of the model (v2.2) by cancer registry and ER status. As expected, for breast cancer specific mortality, v3.0 is well-calibrated in the model development data. It also performs well in the two validation data sets; in all strata of the data the predicted number of deaths was within ten percent of that observed. In contrast, v2.2 consistently over-predicted the number of deaths as might have been expected given the general improvement in prognosis observed since the data on which v2.2 were generated. Prediction of non-breast cancer mortality by v3.0 (Table 4) was also excellent in the model development data, but under predicted by about ten per cent in the validation data sets. Again, v2.2 substantially over predicted other mortality in all the data sets.

**Table 3:** Cumulative observed versus predicted breast cancer deaths estimated by the updated version of PREDICT Breast (v3.0) and the current version (v2.2) by cancer registry and ER status at up to 5, 10 and 15 years follow up.

| Cancer registry |  | No cases | Observed | Predicted |  | Predicted – expected (%) |  |  |  |
| --- | --- | --- | --- | --- | --- | --- | --- | --- | --- |
|  |  |  |  | v3.0 | v2.2 | v3.0 |  | v2.2 |  |
| 5- mortality |  |  |  |  |  |  |  |  |  |
| Eastern | ER + | 5,484 | 908 | 883 | 1,150 | -25 | (-3) | 242 | (27) |
|  | ER - | 34,265 | 1,354 | 1,247 | 1,659 | -107 | (-8) | 305 | (23) |
| West Midlands | ER + | 47,34 | 672 | 642 | 871 | -30 | (-5) | 199 | (30) |
|  | ER - | 27,674 | 900 | 858 | 1,164 | -42 | (-5) | 264 | (29) |
| Others | ER + | 13,369 | 1,643 | 1,560 | 2,377 | -83 | (-5) | 734 | (45) |
|  | ER - | 87,182 | 2,228 | 2,262 | 3,527 | 34 | (2) | 1,299 | (58) |
| 10-year mortality |  |  |  |  |  |  |  |  |  |
| Eastern | ER + | 5,484 | 1,123 | 1,091 | 1,331 | -32 | (-3) | 208 | (19) |
|  | ER - | 34,265 | 2,385 | 2,335 | 2,939 | -51 | (-2) | 554 | (23) |
| West Midlands | ER + | 4,734 | 810 | 807 | 1,022 | -3 | (0) | 212 | (26) |
|  | ER - | 27,674 | 1,509 | 1,600 | 2,040 | 91 | (6) | 531 | (35) |
| Others | ER + | 13,369 | 1,789 | 1,715 | 2,533 | -74 | (-4) | 744 | (42) |
|  |  | 87,182 | 2,865 | 2,963 | 4,472 | 98 | (3) | 1,607 | (56) |
| 15-year mortality |  |  |  |  |  |  |  |  |  |
| Eastern | ER + | 5,484 | 1,155 | 1,120 | 1,349 | -35 | (-3) | 194 | (17) |
|  | ER - | 34,265 | 2,732 | 2,705 | 3,339 | -27 | (1) | 607 | (22) |
| West Midlands | ER + | 4,734 | 826 | 835 | 1,041 | 9 | (1) | 215 | (26) |
|  | ER - | 27,674 | 1,725 | 1,882 | 2,346 | 157 | (9) | 621 | (36) |
| Others | ER + | 13,369 | 1,793 | 1,717 | 2,535 | -76 | (-4) | 742 | (41) |
|  | ER - | 87,182 | 2,890 | 2,983 | 4,494 | 93 | (3) | 1,604 | (55) |

**Table 4:**
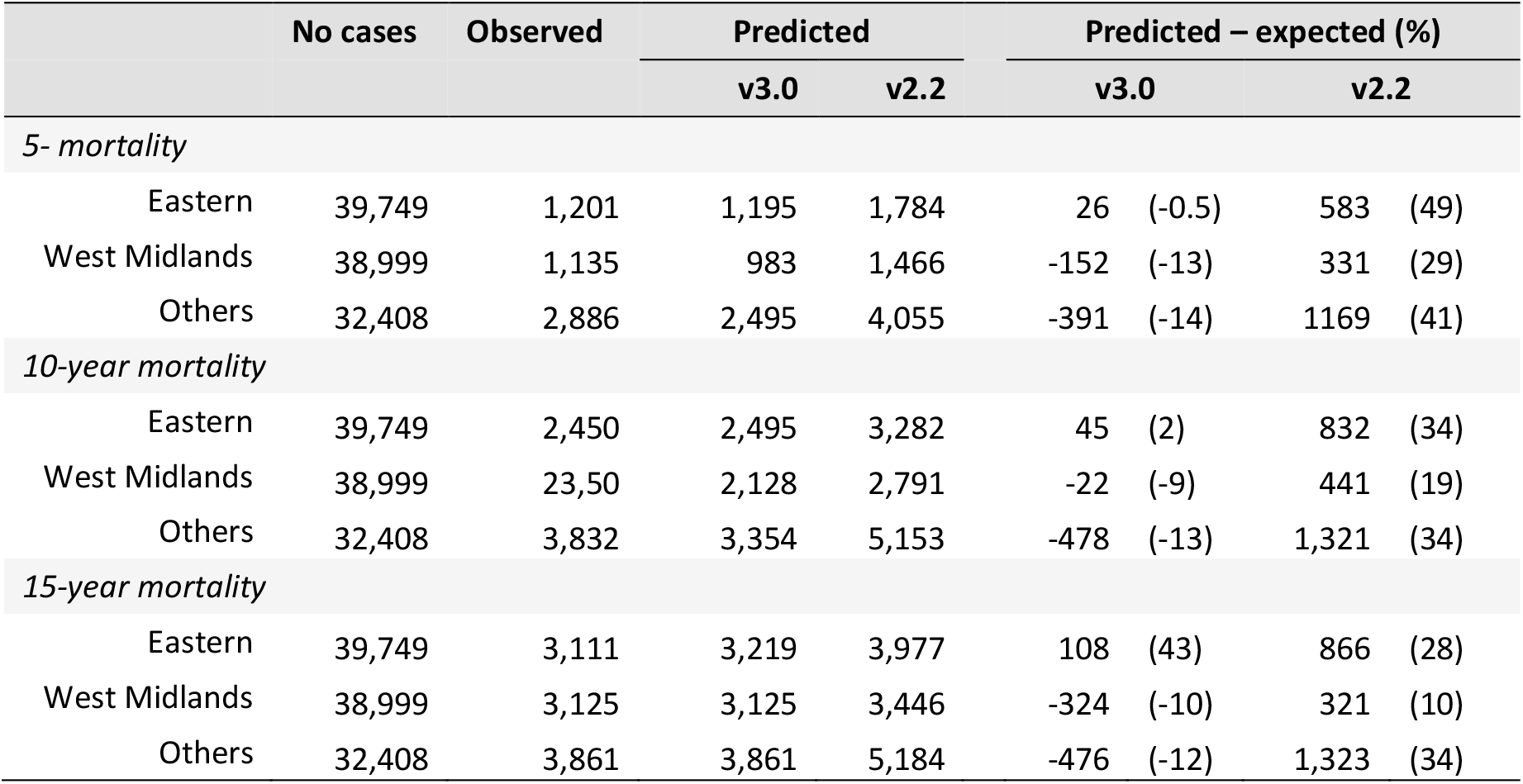
Cumulative observed versus predicted deaths from other causes estimated by the updated version of PREDICT Breast (v3.0) and the current version (v2.2) by cancer registry at up to 5, 10 and 15 years follow up.

**Table 5:** The discrimination for up to 5-year, 10-year and 15-year breast cancer-specific mortality by cancer registry and ER status

| Cancer Registry | ER status | 5-year |  | 10-year |  | 15-year |  |
| --- | --- | --- | --- | --- | --- | --- | --- |
|  |  | v3.0 | v2.2 | v3.0 | v2.2 | v3.0 | v2.2 |
| Eastern Region | ER+ | 0.843 | 0.837 | 0.821 | 0.809 | 0.836 | 0.833 |
|  | ER- | 0.771 | 0.764 | 0.774 | 0.766 | 0.778 | 0.773 |
|  | All | 0.824 | 0.819 | 0.813 | 0.802 | 0.833 | 0.832 |
| West Midlands | ER+ | 0.831 | 0.826 | 0.804 | 0.793 | 0.812 | 0.811 |
|  | ER- | 0.735 | 0.726 | 0.719 | 0.710 | 0.717 | 0.716 |
|  | All | 0.809 | 0.806 | 0.795 | 0.782 | 0.811 | 0.809 |
| Other | ER+ | 0.861 | 0.857 | 0.856 | 0.849 | 0.865 | 0.862 |
|  | ER- | 0.777 | 0.771 | 0.777 | 0.770 | 0.783 | 0.777 |
|  | All | 0.846 | 0.844 | 0.847 | 0.842 | 0.858 | 0.857 |

The observed and predicted breast cancer deaths in the West Midlands cancer registry by quintile of predicted risk for the updated version of PREDICT Breast are shown in Figure 3 which shows that calibration is excellent at all levels of risk.

**Figure 3:**
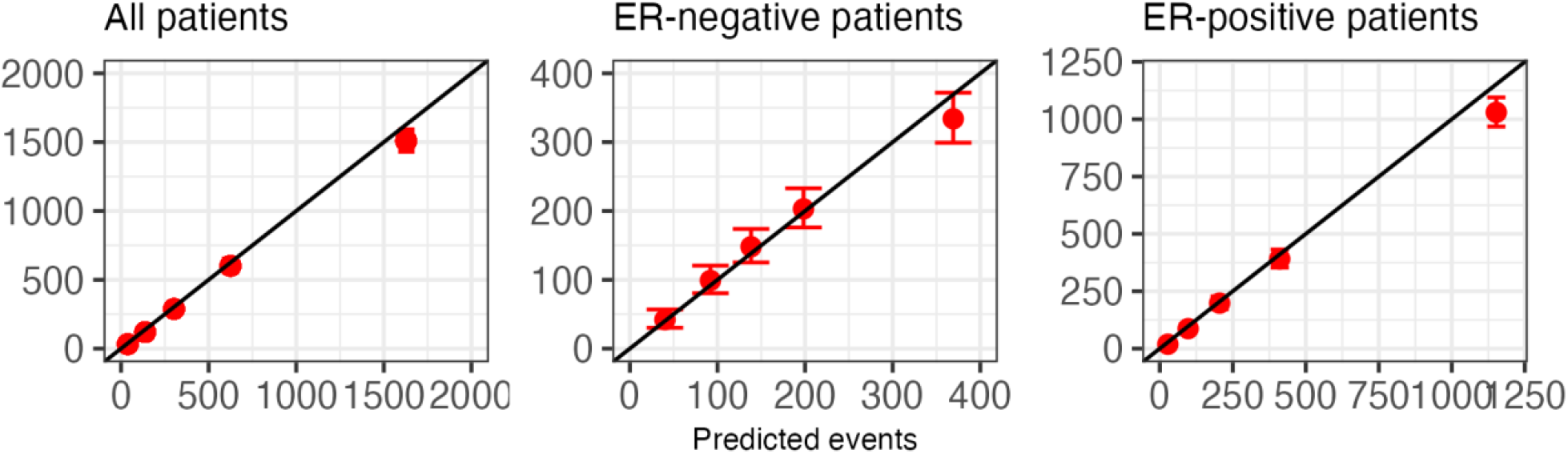
Observed and predicted breast cancer deaths at 15 years in West Midlands data set by quintile of predicted risk for all patients and stratified by ER status.

### Model discrimination

Model discrimination (area under the receiver operator characteristic curve) was good in all strata of the data. In general the model for ER-positive disease performed better than that for ER-negative disease and the performance of the model in the model development data from the Eastern Cancer Registry was slightly better than the performance in the two validation data sets. PREDICT v3.0 performed consistently slightly better than v2.2.

### Model reclassification

The Cambridge Breast Unit classifies women with breast cancer into three groups based on the predicted benefit of adjuvant chemotherapy at 10 years as given by the absolute reduction in risk of breast cancer specific mortality; low-risk women are those with a predicted ten-year benefit of zero to three per cent who would usually be advised not to have adjuvant chemotherapy and high-risk women are those with a predicted benefit of over five per cent who would usually be advised to have adjuvant chemotherapy ^29^. The advice to intermediate risk women (three to five percent) would depend more on other factors including patient preferences. While the benefit of therapy depends on patient age and adjuvant chemotherapy regime it is possible to classify women into similar categories based on the predicted breast cancer mortality at ten years: low risk being zero to fifteen per cent, medium risk being fifteen to twenty per cent and high-risk being greater than 20 per cent risk of breast cancer death at ten years. Based on these risk categories it is possible to evaluate reclassification comparing PREDICT v3.0 with v2.2. Of 32,408 breast cancer cases in the West Midlands data set 4,203 (13%) women would be classified in different risk groups by PREDICT v2.2 and v3.0 (Table 6).

**Table 6:**
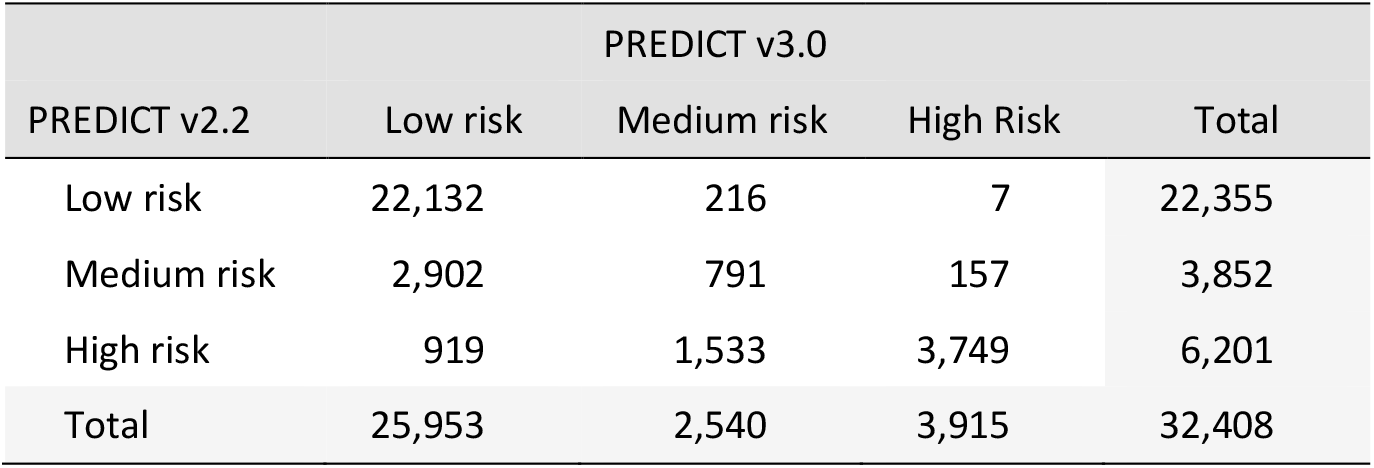
Re-classification of 32,408 West Midlands Cancer Registry breast cancer cases by PREDICT v3.0 into low-, medium- and high-risk compared to PREDICT v2.2 classification.

## DISCUSSION

We have used data from the National Cancer Registration and Analysis Service for England for breast cancer cases diagnosed from 2000 to 2017 to develop and validate a new PREDICT Breast prognostic model (v3.0). We used a similar analytic approach to that used to develop PREDICT Breast v2.0 using multi-variable fractional polynomials within a Cox regression framework to create different models for breast cancer specific mortality for ER-positive disease and ER-negative disease and non-breast cancer mortality. The major difference between v2.2 and v3.0 is that v3.0 includes a term for year of diagnosis as the data show a clear trend from improved survival rates over time.

It has previously been observed that the log hazard ratio function for age at diagnosis in ER-positive breast cancer is U-shaped with breast cancer in young women and older women being associated with a poorer prognosis. However, a similar relationship in ER-negative disease has not been previously described - age at diagnosis in v2.2 was modelled as a linear term. However, in this much larger data set, we also observed a U-shaped function for age at diagnosis in ER-negative disease. We also observed an unexpected hazard ratio function for tumour size in ER-positive cases with an inverted U-shape. There may be a biological reason for this – it is conceivable that for tumours to become very large in size they would need to be growing for a long time without metastasizing, and so may be inherently less aggressive. However, despite our very large data set, the number of ER-positive cases with tumours above 7.5 cm was only 414 with 80 deaths from breast cancer and the precision of the hazard ratio estimates in larger tumours will be small. We therefore chose to constrain the polynomial function for such that the hazard ratio flattened off but did not get smaller with increasing tumour size.

The improvement in prognosis over time is reflected in the reclassification of breast cancer cases within the three categories of risk used by the Cambridge Breast Unit to guide the use of adjuvant chemotherapy. In the West Midlands data set 10,053 cases would be classified as moderate or high risk by PREDICT Breast v2.2 and would be considered candidates for adjuvant chemotherapy. Of these, 3,821 (38%) would be reclassified as low risk by PREDICT Breast v3.0 and spared the harms of chemotherapy.

Tumour gene expression profile tests (also known as genomic risk scores) are being increasingly used to guide treatment decisions in breast cancer ^30^. The results of genomic risk scores are not available in the cancer registration data set used for these analysis and it was not possible to assess any added value of such scores to PREDICT v3.0. However, it has been shown that genomic risk scores do not significantly improve the discrimination of PREDICT v2.2 ^31^. Further research to evaluate the performance of genomic risk scores in breast cancer patients shown to be at intermediate risk by PREDICT v3.0 is warranted.

In an era of precision oncology, accurate, well-validated models that predict patient outcomes are invaluable clinical tools. We have derived an improved version of the PREDICT prognostication and treatment benefit model to reduce some of the limitations of the current version. In particular, we have included updated the model to reflect outcomes in contemporary patients and added the benefits of radiotherapy as well as the harms of both chemotherapy and radiotherapy. The new model has been validated in two independent population-based data sets from the United Kingdom and performs well. It will be implemented in the online tool available at www.breast.predict.nhs.uk and will continue to aid clinical decision making in clinical practice.

## Supporting information

TRIPOD criteria checklist

## DATA AVAILABILITY

The data used for these analyses cannot be shared by the authors for reasons of confidentiality. They are available on request from the England National Disease Registration Service at https://digital.nhs.uk/services/national-disease-registration-service#requests-for-access-to-ndrs-data.

## ACKNOWLEDGEMENTS

We thank: Alex Freeman, David Speigelhalter and Gabriel Recchia for helpful discussion on the development and implementation of the model; and Julia Brown of Public Health England for help in accessing the national cancer registration data set. Isabelle Grootes was funded by the Mark Foundation Institute for Integrated Cancer Medicine at the University of Cambridge.

## SUPPLEMENTARY MATERIAL

**Supplementary Table 1:** Number of breast cancer cases by year of diagnosis and cancer registry with associated data missingness

| Year | No of cases | Complete cases (%) | No cases missing data by variable |  |  |  |  |  |
| --- | --- | --- | --- | --- | --- | --- | --- | --- |
|  |  |  | Any | ER status | Grade | Size | Nodes | Mode of detection |
| Eastern Cancer Registry |  |  |  |  |  |  |  |  |
| 2000 | 1664 | 47 | 888 | 370 | 143 | 87 | 231 | 461 |
| 2001 | 1747 | 49 | 894 | 332 | 123 | 121 | 230 | 514 |
| 2002 | 1946 | 53 | 919 | 303 | 163 | 118 | 262 | 532 |
| 2003 | 2071 | 50 | 1027 | 456 | 156 | 153 | 336 | 510 |
| 2004 | 2196 | 37 | 1377 | 935 | 140 | 165 | 344 | 497 |
| 2005 | 2872 | 34 | 1897 | 1179 | 176 | 208 | 474 | 726 |
| 2006 | 3379 | 41 | 1993 | 1107 | 184 | 216 | 529 | 744 |
| 2007 | 3311 | 58 | 1375 | 336 | 161 | 143 | 468 | 733 |
| 2008 | 3562 | 40 | 2122 | 1690 | 163 | 112 | 435 | 297 |
| 2009 | 3563 | 77 | 803 | 259 | 123 | 156 | 444 | 118 |
| 2010 | 3612 | 82 | 648 | 149 | 96 | 102 | 445 | 74 |
| 2011 | 3499 | 79 | 728 | 101 | 82 | 115 | 556 | 63 |
| 2012 | 3744 | 74 | 972 | 145 | 103 | 102 | 637 | 203 |
| 2013 | 3909 | 74 | 1004 | 159 | 80 | 159 | 728 | 153 |
| 2014 | 3967 | 71 | 1156 | 247 | 85 | 245 | 752 | 167 |
| 2015 | 4073 | 72 | 1125 | 174 | 71 | 233 | 790 | 169 |
| 2016 | 4016 | 67 | 1340 | 277 | 85 | 200 | 894 | 274 |
| 2017 | 3896 | 67 | 1285 | 278 | 47 | 164 | 986 | 83 |
| West Midlands Cancer Registry |  |  |  |  |  |  |  |  |
| 2000 | 1618 | 8 | 1494 | 1486 | 64 | 61 | 43 | 1 |
| 2001 | 1688 | 11 | 1502 | 1486 | 52 | 49 | 38 | 3 |
| 2002 | 1692 | 87 | 228 | 106 | 53 | 48 | 51 | 2 |
| 2003 | 1838 | 88 | 222 | 120 | 51 | 54 | 23 | 2 |
| 2004 | 1930 | 92 | 148 | 75 | 36 | 35 | 23 | 1 |
| 2005 | 2122 | 90 | 209 | 93 | 50 | 65 | 42 | 0 |
| 2006 | 1956 | 91 | 172 | 77 | 45 | 49 | 26 | 1 |
| 2007 | 2073 | 93 | 140 | 49 | 51 | 40 | 22 | 0 |
| 2008 | 2142 | 92 | 168 | 39 | 49 | 74 | 37 | 1 |
| 2009 | 2054 | 91 | 184 | 37 | 46 | 91 | 32 | 1 |
| 2010 | 2246 | 87 | 283 | 90 | 42 | 125 | 63 | 2 |
| 2011 | 3175 | 81 | 611 | 195 | 86 | 228 | 248 | 9 |
| 2012 | 3042 | 74 | 782 | 352 | 66 | 277 | 395 | 14 |
| 2013 | 3272 | 63 | 1209 | 534 | 85 | 361 | 594 | 106 |
| 2014 | 3508 | 63 | 1304 | 506 | 90 | 451 | 669 | 136 |
| 2015 | 3459 | 57 | 1476 | 493 | 91 | 613 | 664 | 229 |
| 2016 | 3399 | 60 | 1343 | 388 | 55 | 410 | 710 | 356 |
| 2017 | 3480 | 59 | 1418 | 561 | 60 | 505 | 809 | 76 |
|  |  |  | Any | ER status | Grade | Size | Nodes | Mode of detection |
| Other Cancer Registries |  |  |  |  |  |  |  |  |
| 2000 | 8496 | 0 | 8469 | 8440 | 651 | 2542 | 6535 | 1171 |
| 2001 | 10112 | 0 | 10068 | 10047 | 693 | 2928 | 6972 | 1671 |
| 2002 | 10840 | 1 | 10785 | 10729 | 638 | 2646 | 6695 | 1873 |
| 2003 | 12712 | 1 | 12643 | 12604 | 681 | 2974 | 7675 | 1841 |
| 2004 | 8509 | 1 | 8436 | 8388 | 410 | 2582 | 6732 | 726 |
| 2005 | 8945 | 1 | 8892 | 8856 | 367 | 2814 | 7159 | 246 |
| 2006 | 8986 | 1 | 8929 | 8901 | 425 | 3173 | 6770 | 137 |
| 2007 | 9010 | 1 | 8951 | 8914 | 335 | 2880 | 7113 | 294 |
| 2008 | 8748 | 1 | 8679 | 8645 | 199 | 1742 | 4045 | 178 |
| 2009 | 9545 | 4 | 9170 | 9079 | 234 | 664 | 2552 | 73 |
| 2010 | 10359 | 35 | 6776 | 6186 | 245 | 1101 | 2521 | 95 |
| 2011 | 15079 | 66 | 5109 | 3308 | 360 | 1607 | 2842 | 185 |
| 2012 | 22439 | 61 | 8803 | 5290 | 708 | 3286 | 4544 | 351 |
| 2013 | 23117 | 62 | 8764 | 4233 | 437 | 3305 | 4302 | 1020 |
| 2014 | 25231 | 56 | 11010 | 5551 | 550 | 4463 | 5729 | 1202 |
| 2015 | 25926 | 53 | 12107 | 5858 | 397 | 5534 | 5511 | 1509 |
| 2016 | 26171 | 47 | 13753 | 7718 | 337 | 4271 | 5563 | 3132 |
| 2017 | 26164 | 50 | 13096 | 7956 | 360 | 4237 | 6485 | 866 |

**Supplementary Figure 1:**
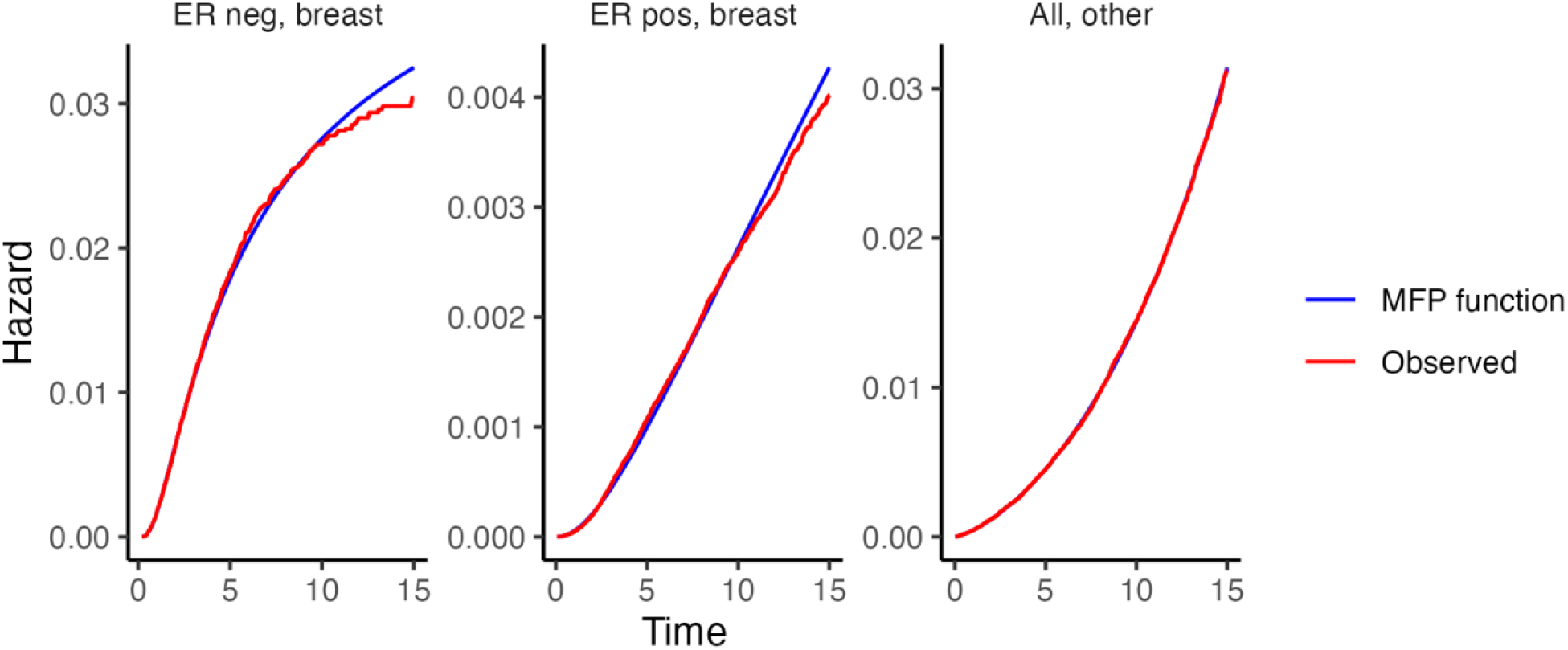
Observed baseline hazard and fitted polynomial baseline hazard function for ER-positive breast cancer specific mortality, ER-negative breast cancer specific mortality and non-breast cancer mortality

